# Prevalence, Genotyping, and Mutational Analysis of Hepatitis B Virus in HIV-Infected Patients on Antiretroviral Therapy in Nairobi, Kenya

**DOI:** 10.64898/2026.05.06.26352536

**Authors:** Lydia Ochieng, Rosaline Macharia, Matilu Mwau

**Affiliations:** Department of Biochemistry, University of Nairobi, Nairobi, Kenya; Kenya Medical Research Institute, Nairobi, Kenya

**Keywords:** Hepatitis B virus, Human immunodeficiency virus, HIV/HBV coinfection, HBV genotypes, Escape mutations, Kenya

## Abstract

**Background:** Hepatitis B virus infection remains a major public health challenge, particularly among people living with human immunodeficiency virus, due to shared transmission routes and the potential for accelerated liver disease progression. Molecular characterization of circulating HBV strains is essential for understanding viral epidemiology, mutation patterns, and implications for diagnostics and vaccination.

**Methods:** This study investigated the prevalence of hepatitis B infection and molecular characteristics of the hepatitis B virus surface gene among HIV-infected individuals receiving antiretroviral therapy in Nairobi County, Kenya. Plasma samples were screened for hepatitis B surface antigen using enzyme-linked immunosorbent assay. Hepatitis B viral DNA was extracted from HBsAg-positive samples and the surface gene region amplified by polymerase chain reaction. Amplified products were subjected to Sanger sequencing. Sequence assembly, genotype determination, and mutation analysis.

**Results:** The prevalence of HIV/HBV co-infection among HIV-positive individuals was determined to be 8.97%. Genotype analysis revealed the circulation of HBV genotype A (sub-genotypes A1 and A4) and genotype D (sub-genotypes D4 and D10) among the studied population. Amino acid sequence analysis of the major hydrophilic region of the surface gene identified several mutations, with R122K and Y134F being the most frequently observed substitutions.

**Conclusion:** Hepatitis B infection remains prevalent among HIV-infected individuals receiving antiretroviral therapy in Nairobi County. The circulation of multiple hepatitis B virus genotypes and the presence of mutations within the surface gene highlight the importance of continuous molecular surveillance to monitor viral evolution and its potential implications for hepatitis B virus diagnosis, vaccination strategies, and clinical management in HIV-infected populations

## Introduction

Hepatitis B virus (HBV) infection continues to be a significant global public health concern, with an estimated 296 million people living with chronic infection worldwide (WHO, 2021). This condition is responsible for approximately 820,000 deaths each year, primarily due to complications such as liver cirrhosis and hepatocellular carcinoma (HCC) (Flores *et al*., 2022; WHO, 2021). The burden of HBV is particularly high in sub-Saharan Africa (SSA), where endemic transmission plays a crucial role in liver-related illnesses and deaths (Sheena *et al*., 2022; WHO, 2022). In Africa, the prevalence of chronic HBV infection among adults is estimated at 6.1% (Makuza J *et al*., 2019), with several countries, including Burkina Faso, Nigeria, Zimbabwe, Mali, Ghana, and Gabon classified as hyper-endemic, reporting hepatitis B surface antigen (HBsAg) prevalence rates exceeding 8% (McNaughton *et al*., 2020).

Human immunodeficiency virus (HIV) infection often overlaps geographically with HBV, primarily due to shared transmission routes such as sexual contact, exposure to infected blood, and perinatal transmission (Cheng *et al*., 2021). Sub-Saharan Africa, especially East and Southern Africa, bears the highest global burden of HIV infection, with approximately 20.7 million people living with HIV as of 2019 (Parker *et al*., 2021; UNAIDS, 2020). This overlap results in a notable number of individuals living with HIV/HBV co-infection. In Kenya, HBV is endemic, with reported prevalence rates ranging from 2to 8% in the general population (Karoney *et al*., 2020). Higher rates are noted among high-risk groups, particularly individuals living with HIV (Kerubo *et al*., 2015; Makokha *et al*., 2023; Muriuki *et al*., 2013). Studies among HIV-infected individuals in Kenya has shown that the prevalence of HIV/HBV co-infection ranges from approximately 4% to over 10%. This variation depends on geographic, demographic, and methodological factors (Karoney *et al*., 2020; Kirui, 2018; Maina *et al*., 2017).

Human immunodeficiency virus and HBV co-infection has significant clinical implications. In individuals co-infected with HIV, HBV replication can be enhanced due to HIV-related immunosuppression, leading to higher HBV viral loads, delayed or reduced rates of spontaneous HBeAg seroconversion, and accelerated progression to chronic liver disease, cirrhosis, and HCC (Mokaya *et al*., 2018). As antiretroviral therapy (ART) has expanded across SSA, HIV-related morbidity and mortality have significantly declined, resulting in improved survival and increased life expectancy among individuals living with HIV (Klein *et al*., 2011; Wong *et al*., 2019). However, as these individuals age, chronic comorbidities like liver disease have emerged as major contributors to non-AIDS-related mortality (Chambal *et al*., 2017; Wan *et al*., 2022). Despite the widespread use of ART, liver disease remains a leading cause of death among those living with HIV, highlighting the urgent need for comprehensive HBV screening, molecular monitoring, and optimized management within HIV care programs.

Beyond its epidemiological burden, HBV is characterized by substantial genetic variability. The HBV genome consists of a small, partially double-stranded DNA of approximately 3.2 kb organized into four overlapping open reading frames (Surface, Core, Polymerase, and X), which encode surface (HBsAg), core (HBcAg), polymerase, and regulatory X proteins, respectively (Schollmeier *et al*., 2023). Due to the error-prone nature of the reverse transcription process during replication, HBV accumulates mutations at a relatively high rate, contributing to its genetic diversity (Liu *et al*., 2020).

Based on sequence divergence across the complete genome, HBV is classified into at least 10genotypes (A–J), which differ in geographical distribution, transmission dynamics, disease progression patterns, and response to therapy (Chen *et al*., 2023). In Africa, genotypes A, D, and E are most commonly reported, with genotype A predominating in Eastern and Southern Africa (Mabeya *et al*., 2016; Nyairo, 2018).

Molecular variations within the HBV genome, particularly in the surface (S) gene that encodes HBsAg, are clinically significant (Shoraka *et al*., 2023). Mutations within or surrounding the Major Hydrophilic Region (MHR) at amino acid positions 99 to 169, specifically in the “a” determinant region (amino acids 124 to 147), may alter antigenicity, leading to immune escape, reduced detection by diagnostic assays, vaccine escape, and potential failure of hepatitis B immunoglobulin therapy (Lazarevic *et al*., 2019; Sun *et al*., 2018). Some mutations have also been associated with occult HBV infection and HCC (Al-Qahtani *et al*., 2017).

In Kenya, several studies have documented the presence of HBV genotypes and S gene mutations, including immune-escape variants, across different populations (Aluora *et al*., 2020; Kerubo *et al*., 2015; Mabeya *et al*., 2016; Nyairo, 2018). However, information on the molecular characteristics of HBV is insufficient, more so for HBsAg mutants circulating among HIV-infected patients. Also, information on the impact of ART treatment on HBV during HIV/HBV co-infection is still elusive (Cheng *et al*., 2021). Understanding the circulating genotypes and mutation patterns within this group is critical for informing public health strategies, improving diagnostic sensitivity, optimizing vaccine effectiveness, and guiding antiviral therapy.

This study aimed to determine the prevalence of HIV/HBV co-infection in Nairobi County and to characterize the molecular profiles of HBV strains circulating among HIV-positive individuals. Specifically, we sought to identify HBV genotypes and analyze mutations within the S gene, with emphasis on clinically significant variants associated with immune escape, diagnostic failure, and vaccine escape. The findings will contribute to ongoing molecular surveillance efforts and provide valuable insights for HBV management within HIV care programs in Kenya.

## Materials and Methods

### Study Design and Setting

This was a cross-sectional laboratory-based study conducted using archived remnant EDTA plasma samples obtained from HIV-positive individuals undergoing routine viral load monitoring. The samples were from 10 sub-county hospitals in Nairobi County: Langata, Dagoretti, Starehe, Ruaraka, Kasarani, Embakasi West, Kamukunji, Embakasi East, Westlands, and Makadara. These hospitals serve as referral hubs for approximately 50 affiliated public health facilities catering to both urban and peri-urban populations within the county. Nairobi County, the capital and largest metropolitan area in Kenya, has an estimated population of around 4.3 million people and carries a significant burden of HIV infection. The county hosts an extensive network of Comprehensive Care Clinics (CCCs) that provide ART, routine laboratory monitoring, and long-term HIV care to a diverse patient population. The centralized referral of specimens to the Kenya Medical Research Institute (KEMRI) laboratory, which processes approximately 500 plasma samples daily for HIV viral load testing, ensures a broad representation of HIV-infected individuals receiving care throughout the county. This setting thus provides an appropriate and epidemiologically relevant population for assessing the prevalence and molecular characteristics of HBV among people living with HIV. Sample selection and all laboratory analyses were conducted at the KEMRI Laboratory for Molecular Biology in Nairobi.

### Study Population

This study comprised HIV-positive adults receiving care at CCCs in Nairobi County for routine HIV treatment monitoring. Eligible participants were individuals aged 18 years or older who had been on ART for at least six months at the time of sample collection, regardless of gender. Samples were collected from participants without considering their prior knowledge of HBsAg status.

### Sample Size

A total of 870 HIV-positive study participants was deemed sufficient to estimate the prevalence of HIV/HBV co-infection, using a significance level of 0.05. The sample size was calculated using Cochran’s (1963) formula for cross-sectional studies, based on an estimated co-infection prevalence of 6% from previous research conducted in Kenya (Muriuki *et al*., 2013). This calculation indicated that a minimum of approximately 87 participants was required from each of the 10 sub-county hospitals, resulting in a total sample size of 870 participants.

### Sample Selection and Processing

Following completion of routine HIV viral load testing, plasma samples were stored at −20°C in the KEMRI Laboratory for Molecular Biology. For this study, a total of 870 archived remnant samples collected in K₂EDTA vacutainer tubes were retrieved from storage. From each sample, 600 µL of plasma was aliquoted into 1.5 mL cryovials labeled with unique study identification numbers and arranged in racks for storage. The aliquots were maintained at −20°C until further laboratory analysis.

### Serological Screening for HBV Infection

All 870 samples were screened for HBsAg using BioELISA HBsAg 3.0 Biokit assay following the manufacturer’s manual (Biokit., 2018). Samples that tested positive for HBsAg were considered indicative of current HBV infection and were subjected to molecular analysis.

### Hepatitis B Viral Load Quantitation

All HBsAg-positive plasma samples were subjected to quantitative HBV DNA analysis. A total of 400 µL of plasma was used for viral DNA extraction and amplification, following the manufacturer’s instructions for the Abbott RealTime HBV assay (Abbott Molecular, USA). The assay was conducted on the Abbott m2000 system, which quantifies HBV DNA using real-time polymerase chain reaction (PCR). Samples with HBV DNA levels exceeding 1,000 copies/mL were considered indicative of active viral replication and were selected for further amplification and sequencing of the HBV S gene.

### Amplification of the HBV S Gene for Sequencing

The HBV S gene region spanning nucleotides 231 to 801 was amplified using a nested PCR approach on an ABI Veriti™ 96-well thermal cycler (Applied Biosystems, USA).

The first-round PCR was performed using forward primer PS1-F (5′-TCACAATACCGCAGAGTCT-3′) and reverse primer PS1-R (5′-AACAGCGGTATAAAGGGACT-3′), targeting nucleotides 1 to 571 base pairs of the S gene. Each 50 µL PCR reaction mixture comprised 25 µL of HotStarTaq Master Mix (Qiagen, Hilden, Germany), 5 µL each of 10 µM PS1-F and PS1-R primers, 5 µL of template DNA, and nuclease-free water to volume. Thermal cycling conditions included an initial denaturation at 94°C for 3 minutes, followed by 40 cycles of denaturation at 94°C for 30 seconds, annealing at 55°C for 30 seconds, and extension at 72°C for 1 minute, with a final extension step at 72°C for 5 minutes.

The second-round amplification employed primers PS2-F (5′-GTGGTGGACTTCTCTCAATTTTC-3′) and PS2-R (5′-CGGTATAAAGGGACTCACGAT-3′), yielding an expected amplicon size of 570 base pairs. The second-round PCR was performed in a total reaction volume of 50 µL under identical cycling conditions as the first round, except that the number of amplification cycles was reduced to 30.

The generated amplicons were analyzed using electrophoresis on a 1% agarose gel prepared with 1X Tris-acetate-EDTA (TAE) buffer. A total of 5μL of the amplicons were loaded onto the gel and electrophoresed at 100 V for an hour. The DNA bands were then visualized under ultraviolet (UV) transillumination using a Clearview Ultra Violet Transilluminator (Cleaver Scientific Ltd, Rugby, UK). Fragment sizes were determined by comparing the bands to a 1000 bp DNA ladder (Qiagen, Hilden, Germany), with the HBV S gene expected to be about 570 bp. Gel images were captured using a digital documentation system. The nested PCR products were purified for sequencing using the Exonuclease I/Shrimp Alkaline Phosphatase (EXO-SAP) cleanup reagent (Affymetrix, Inc., USA), following a well-established protocol (Appliedbiosystems, 2019).

### DNA Sequencing

Bidirectional Sanger sequencing of the amplified HBV S gene fragments was performed using the same primer pairs employed during PCR amplification. Sequencing reactions were prepared using the BigDye™ Terminator v3.1 Cycle Sequencing Kit (Thermo Fisher Scientific, Waltham, MA, USA) according to the manufacturer’s protocol. Each reaction contained 20 µL of BigDye Terminator v3.1 reaction mixture added to the PCR products. Cycle sequencing was performed for 25 cycles consisting of denaturation at 96 °C for 30 seconds, annealing at 50 °C for 5 seconds, and extension at 60 °C for 4 minutes. Sequencing products were purified using the BigDye XTerminator™ Purification Kit (Thermo Fisher Scientific, Waltham, MA, USA) according to the manufacturer’s instructions. The purified products were analyzed in the ABI 3730XL DNA Analyzer (Applied Biosystems, USA) to generate nucleotide sequences of the HBV S gene amplicons.

### Sequence Assembly, Genotyping and Mutation Analysis of HBV S gene

Raw sequence chromatograms were visually inspected for quality, and ambiguous base calls were edited before analysis. Forward and reverse sequences were assembled into consensus sequences using BioEdit version 7.2.5 (Hall, 1999). The resulting consensus sequences were aligned with 17 HBV sequences retrieved from the National Center for Biotechnology Information (NCBI) GenBank database using the Multiple Sequence Comparison by Log-Expectation (MUSCLE) algorithm implemented in Molecular Evolutionary Genetics Analysis (MEGA) version 10.2.5 (Edgar, 2004; Kumar *et al*., 2018).

The HBV genotypes were determined through phylogenetic analysis based on the amplified S gene region. Multiple sequence alignment was performed using the MUSCLE algorithm implemented in MEGA version 10.2.5 against representative sequences of known HBV genotypes obtained from GenBank. Phylogenetic trees were constructed using the Maximum Likelihood method with the Kimura two-parameter model to infer evolutionary relationships (Kimura, 1980). The robustness of the phylogenetic tree topology was assessed by bootstrap analysis with 1,000 replicates (Felsenstein, 1985). Genotype assignment was based on clustering patterns with reference sequences of known HBV genotypes. Genotyping results were further confirmed using the NCBI HBV genotyping tool (Rozanov *et al*., 2004).

Mutation analysis of the HBV S gene was conducted by comparing the study sequences with genotype-specific reference sequences using MEGA version 10.2.5 (Kumar *et al*., 2018). Particular attention was given to amino acid substitutions within the surface gene that have previously been associated with immune escape, vaccine escape, and diagnostic detection failure. Observed variation patterns were interpreted based on previously published reports (Ababneh *et al*., 2019; Henock et al., 2018; Nyairo, 2018). Identified mutations were further verified using the Geno2pheno [HBV] v2.0 web-based tool (https://hbv.geno2pheno.org).

The 34 HBV S gene nucleotide sequences generated in this study were submitted to the NCBI GenBank database under accession numbers PZ062441-PZ062474. The accession numbers corresponding to each study sample are provided in **Error! Reference source not found.**.

### Data Management and Statistical Analysis

Laboratory results, including HBsAg ELISA outcomes and HBV viral load measurements, were recorded and managed using Microsoft Excel (Microsoft Corporation, USA). Statistical analyses were performed using Stata/SE version 13.0 (StataCorp LP, College Station, TX, USA). Descriptive statistics were used to summarize the study findings. The prevalence of HBV infection was calculated as the proportion of samples that tested positive for HBsAg relative to the total number of samples tested and was expressed as a percentage with 95% confidence intervals.

### Ethical considerations

The Kenya Medical Research Institutés Scientific and Ethics Review Unit (SERU) reviewed and approved this study under KEMRI/SERU Protocol No. 3942.

## Results

### Prevalence of HBV infection among HIV-positive individuals

A total of 870 archived plasma samples obtained from HIV-positive individuals receiving care in Nairobi County were screened for HBsAg using ELISA. Among these, 78 samples tested positive, while 792 samples were negative for HBsAg. The overall prevalence of HBV infection among HIV-positive individuals was determined to be 8.97% (78/870; 95% CI: 7.24–11.06%) (Table 1).

**Table 1:**
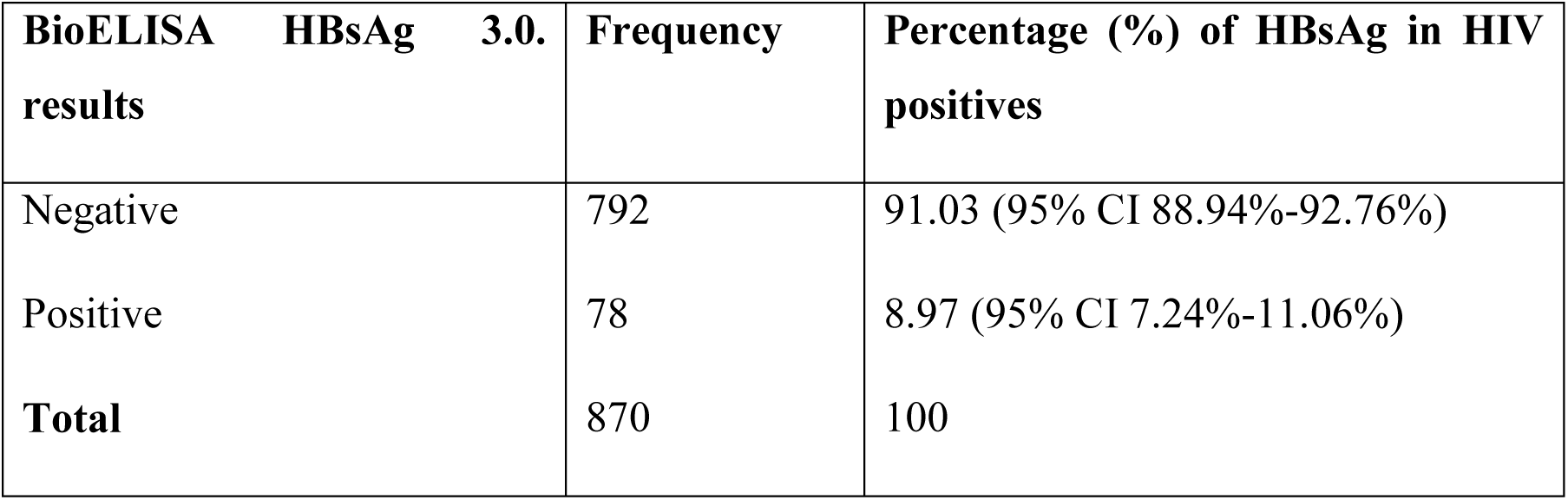
Hepatitis B prevalence in HIV positive patients in Nairobi County.

### Active HBV infection in HBV/HIV co-infection

All 78 HBsAg-positive plasma samples were further tested for HBV DNA using the Abbott Real-Time HBV viral load assay. Hepatitis B viral DNA was detected in 38 samples (48.72%), indicating active viral replication among nearly half of the seropositive individuals. The quantified HBV DNA levels varied among the positive samples, with viral load values summarized using descriptive statistics. Based on the study criteria for active infection, 34 samples had HBV DNA levels exceeding 1000 copies/mL and were therefore considered eligible for subsequent amplification and sequencing of the HBV S gene.

Overall, the proportion of HIV-infected individuals with active HBV infection (HBsAg positive with detectable HBV DNA >1000 copies/mL) in the study population was 3.91% (34/870).

### Amplification and Sequencing of the HBV S Gene

Plasma samples with HBV DNA levels exceeding 1000 copies/mL (n = 34) were subjected to nested PCR amplification targeting the HBV S gene region. The amplification produced the expected 570bp amplicon, which was subsequently purified for downstream sequencing analysis. The purified PCR products were sequenced using the Sanger sequencing method, generating both forward and reverse sequence reads for each sample to enable accurate sequence assembly and molecular characterization of the HBV S gene.

### HBV Genotype Distribution among HIV/HBV Co-infected Individuals

Phylogenetic analysis of the HBV S gene sequences was performed to determine the genotype distribution among the study samples. A total of 34 sequences obtained from successfully amplified samples were included in the analysis and aligned with reference HBV sequences representing known genotypes retrieved from GenBank.

The phylogenetic analysis revealed the presence of two HBV genotypes, namely genotype D and genotype A, circulating among HIV/HBV co-infected individuals in Nairobi County. Genotype D was the predominant genotype, identified in 26 of the 34 sequences (76.47%), while genotype A was detected in 8 sequences (23.53%).

The evolutionary relationships between the study sequences and reference genotype sequences are illustrated in the phylogenetic tree (Figure 1).

**Figure 1:**
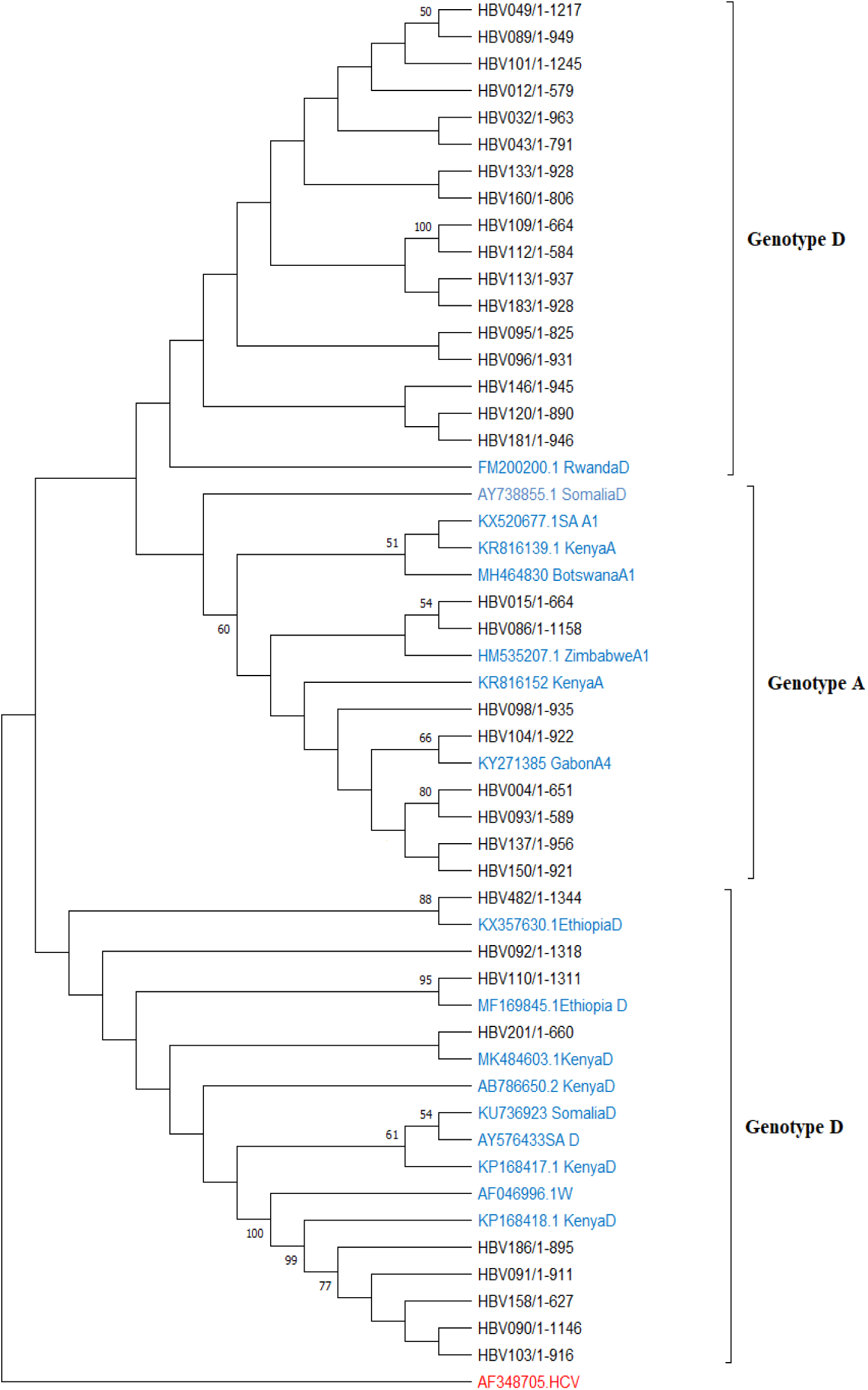
Phylogenetic tree of HBsAg gene sequences generated using the Maximum Likelihood (ML) statistical method from 34 HBV isolates. Each isolate has the prefix ‘HBV’. The isolates were compared with 17 other sequences retrieved from GenBank. The accession numbers of the downloaded sequences followed by country of origin and their HBV genotype are provided (marked in blue). The figures at the nodes indicate the percentage of bootstrap values. Only the values ≥50% were displayed following the bootstrap analysis to confirm the relevance of the phylogenetic tree assessment. The tree was rooted using the Hepatitis C virus (HCV) sequence (marked in red).

### Mutation Analysis of the HBV S Gene

Mutation analysis of the HBV S gene sequences was performed to identify amino acid substitutions associated with immune escape, diagnostic detection failure, and vaccine escape. Comparison of the study sequences with genotype-specific reference sequences revealed several mutations within the surface gene region.

Among the identified mutations, R122K, N143S, and V168A were detected and have previously been associated with diagnostic escape, potentially affecting the detection of hepatitis B surface antigen by certain diagnostic assays. In addition, several mutations linked to vaccine escape were observed, including T114S, Y134F, G145V, N146K, T148P, and E164G.

These mutations occurred at varying frequencies among the analyzed sequences. A summary of the identified amino acid substitutions and their reported clinical significance is presented in Table 2.

**Table 2:**
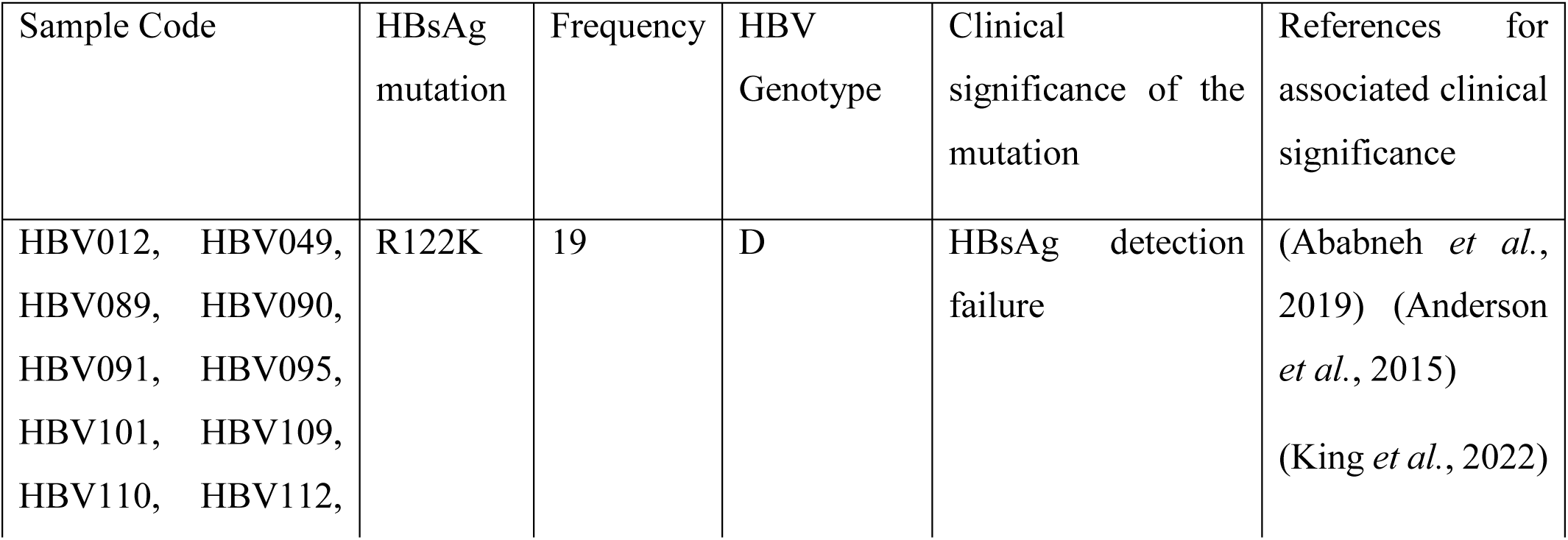
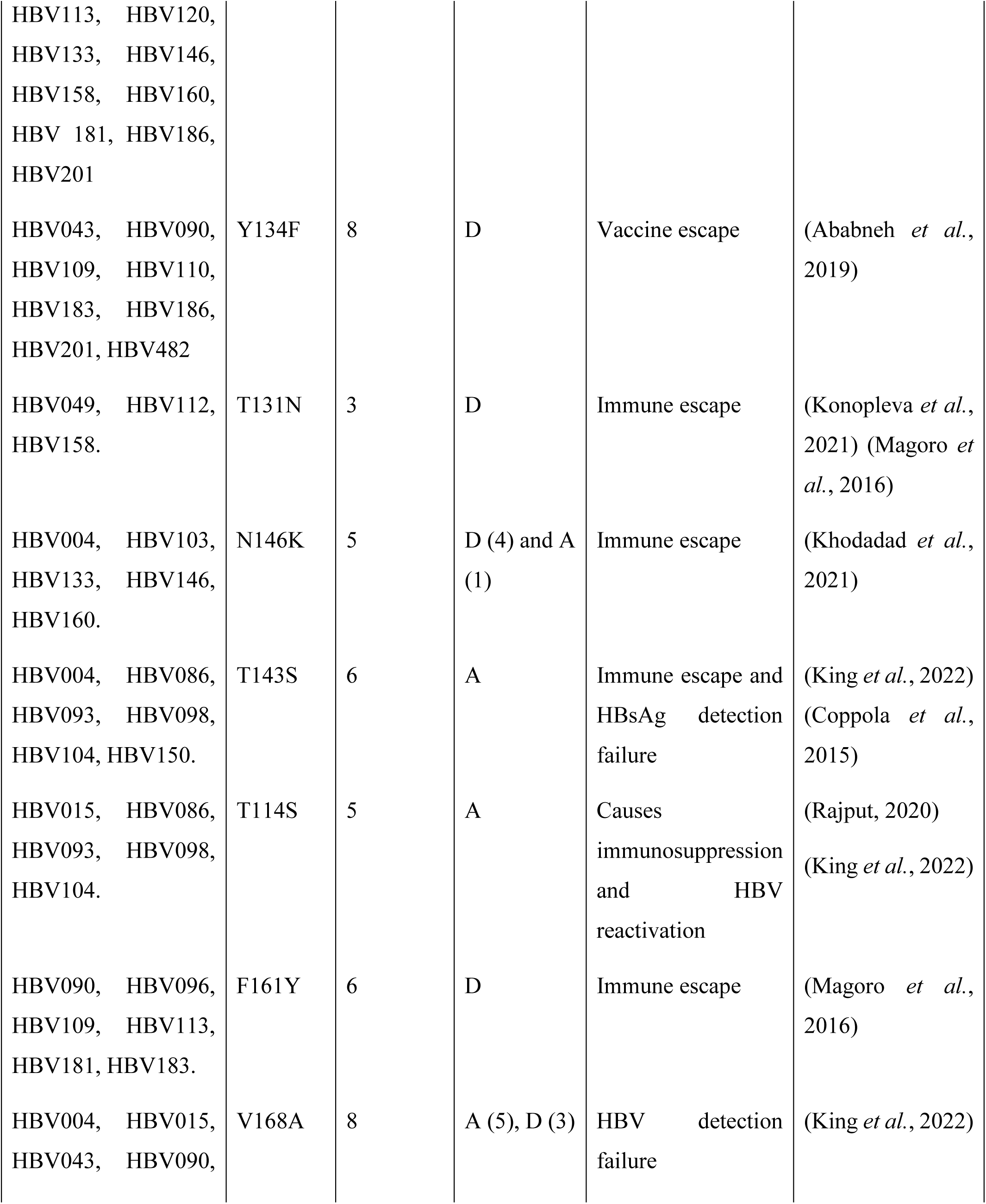
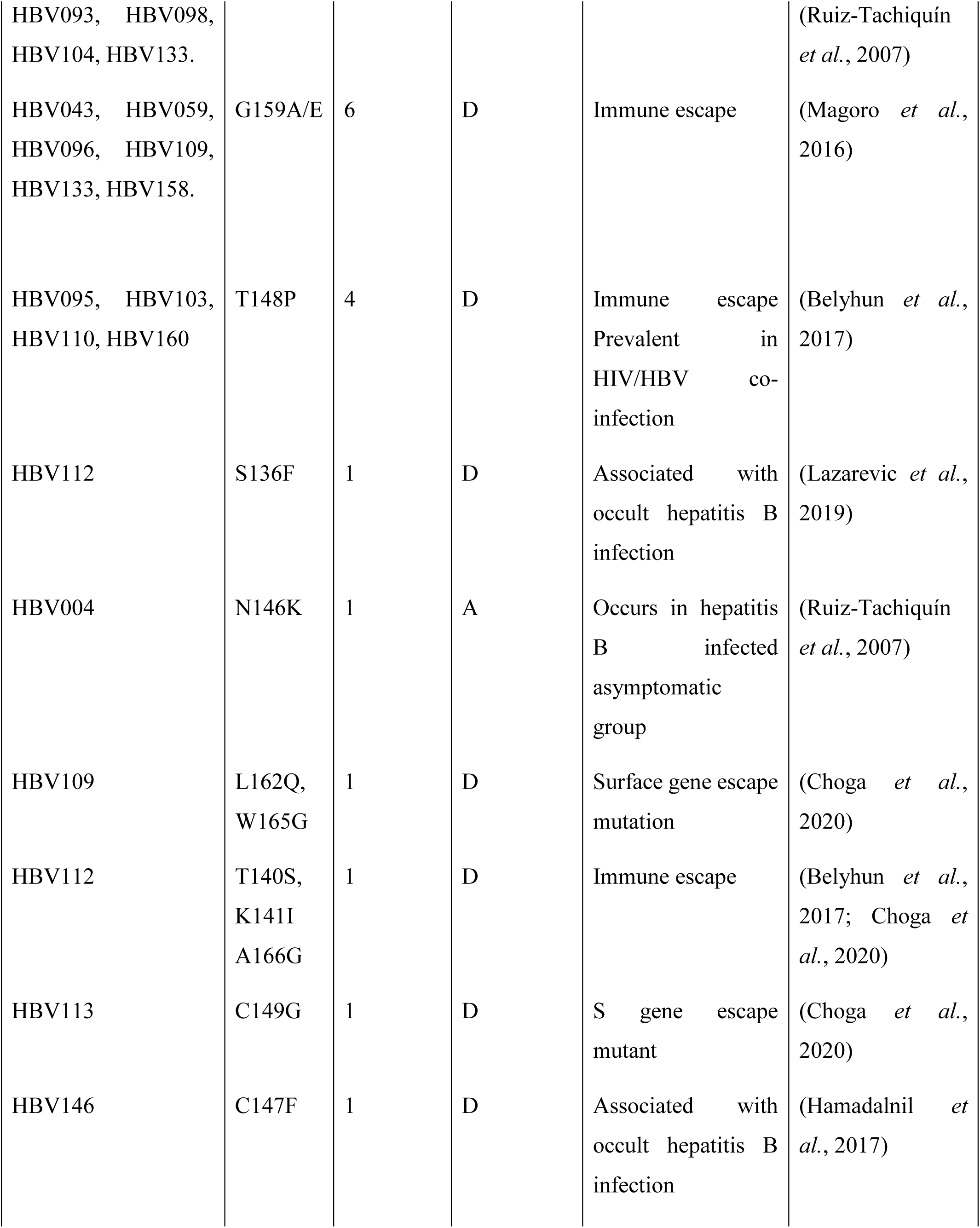
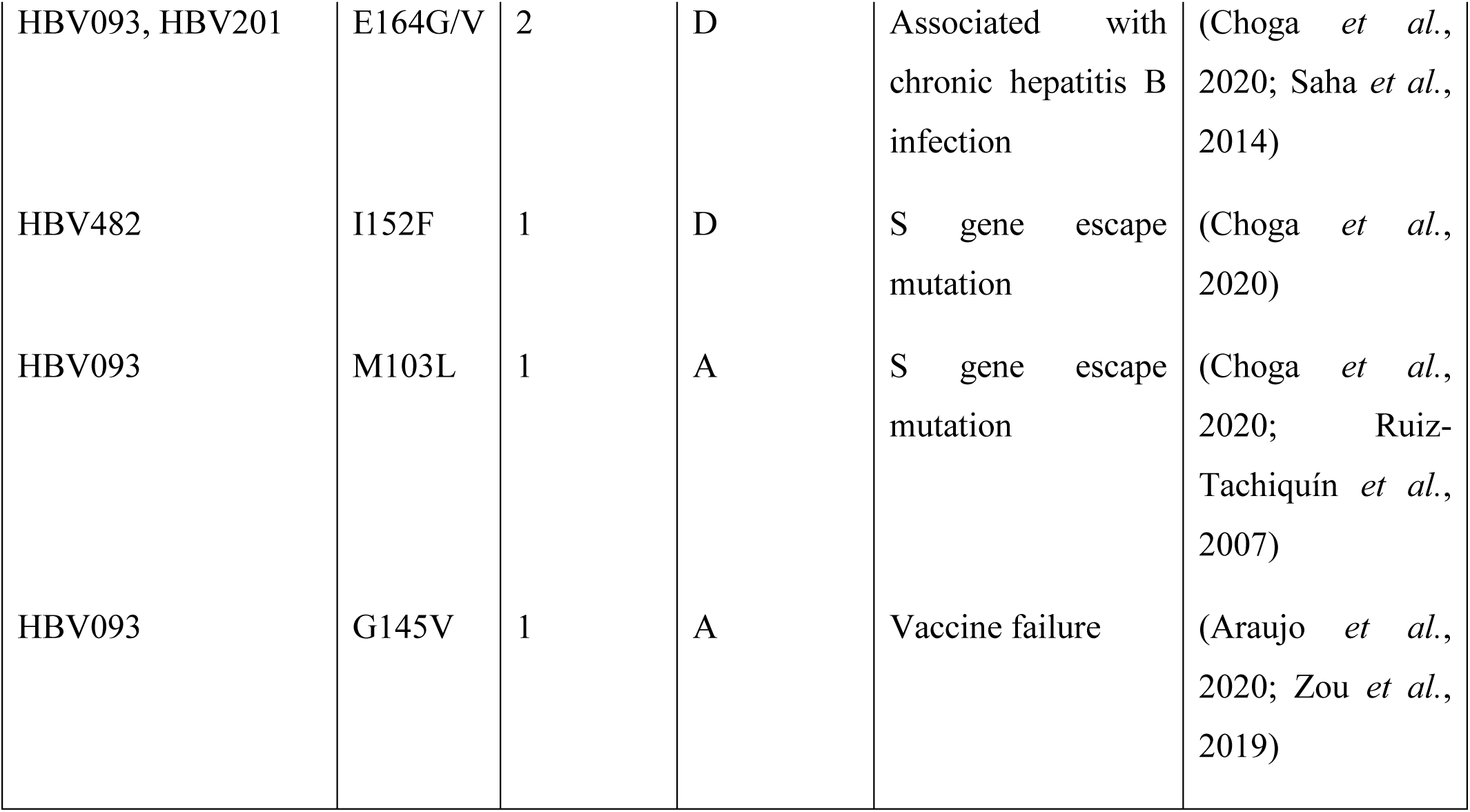
The HBsAg mutations detected within the MHR of the HBV sequences isolated from HBV/HIV co-infected individuals under ART between 2018 and 2019 in Nairobi, Kenya.

## Discussion

This study investigated the prevalence and molecular characteristics of the HBV among individuals infected with HIV who are receiving care in Nairobi County, Kenya. The findings revealed that the prevalence of HBV among HIV-positive individuals is 8.97%, highlighting a significant burden of co-infection within this population. This prevalence is comparable to previous reports from Kenya, where the prevalence of HBV in people living with HIV has been reported to range between 4% and 10%, depending on the study population and diagnostic methods used (Karoney *et al*., 2020; Kirui, 2018; Mabeya *et al*., 2016; Maina *et al*., 2017; Muriuki *et al*., 2013). Similar prevalence estimates have also been observed in other African countries, including Gabon, Mozambique, and Nigeria, which reflects the ongoing epidemiological overlap between HBV and HIV infections in these regions (Bivigou-Mboumba *et al*., 2018; Chambal *et al*., 2017; Karoney *et al*., 2020; Lesi *et al*., 2007). The similarity in prevalence rates can be explained by the fact that these studies were conducted among people living with HIV/AIDS (PLWHA). The observed burden of co-infection may be attributed to shared routes of transmission for HBV and HIV, such as sexual contact, exposure to infected blood, and perinatal transmission, which facilitate the concurrent spread of these infections in areas with high HIV prevalence (Cheng *et al*., 2021; WHO, 2021).

In addition, HBV DNA was detected in 48.72% of samples that tested positive for HBsAg, indicating active viral replication in nearly half of individuals with serologically confirmed HBV infection. Among these, 34 samples had HBV DNA levels exceeding 1,000 copies/mL, which represents 3.91% of the total study population with active HBV infection. The presence of HBV DNA in HBsAg-positive individuals highlights the significance of molecular assays in identifying ongoing viral replication and assessing the clinical activity of HBV infection. Similar findings have been reported in other studies involving HIV-infected populations, where a considerable proportion of HBsAg-positive individuals were found to have detectable HBV DNA (Mokaya *et al*., 2018; Ochwoto *et al*., 2016). Research indicates that HIV-associated immunosuppression can enhance HBV replication, resulting in higher viral loads and reduced rates of spontaneous viral clearance among co-infected individuals (Choi *et al*., 2019; WHO, 2022). These findings emphasize the necessity for routine HBV screening and viral load monitoring in HIV care programs to ensure early identification and management of active HBV infection.

Phylogenetic analysis of the HBV S gene sequences revealed the circulation of two HBV genotypes, genotype D and genotype A, among HIV/HBV co-infected individuals in Nairobi County. Genotype D was the predominant type, accounting for 76.47% of the sequences, while genotype A represented 23.53%. This predominance of genotype D aligns with findings from several studies conducted in Kenya and other parts of Africa, where genotypes A and D are commonly reported among HIV-infected populations (Hamida *et al*., 2021; King *et al*., 2022; Patel *et al*., 2020). Previous molecular epidemiological studies in Kenya have also documented the co-circulation of these genotypes, with genotype A historically recognized as the dominant genotype in East Africa (Kramvis, 2018; Nyairo, 2018; Ochwoto *et al*., 2016). The identification of both genotypes in this study underscores the genetic diversity of HBV circulating in the region, which may reflect the dynamic transmission patterns associated with urban populations like Nairobi, where significant population movement and interaction occur. Differences in HBV genotype distribution are clinically relevant, as certain genotypes have been linked to variations in disease progression, treatment response, and mutation patterns (Cheng *et al*., 2021; Fletcher *et al*., 2020).

The mutation analysis of the HBV S gene sequences identified several amino acid substitutions within the surface gene, including mutations previously associated with diagnostic escape and vaccine escape. Among these, R122K, N143S, and V168A have been reported to influence the antigenic properties of the HBsAg and may potentially affect the sensitivity of certain diagnostic assays used for HBV detection (Ababneh *et al*., 2019; Ambachew *et al*., 2018). In addition, mutations such as T114S, Y134F, G145V, N146K, T148P, and E164G have been described in previous studies as being associated with vaccine escape variants, particularly when they occur within or near the MHR of the HBV surface antigen (Hosseini *et al*., 2019; Ochwoto *et al*., 2016).The emergence of such mutations is of clinical and public health importance because they may alter antigenicity and potentially reduce the effectiveness of immune responses generated by vaccination or immunoglobulin therapy (Magoro *et al*., 2016). Similar mutations within the S gene have been reported in HBV molecular surveillance studies conducted in Africa and other regions, emphasizing the importance of continuous monitoring of circulating HBV strains to inform vaccine strategies and improve diagnostic tools (Ababneh *et al*., 2019; Mokaya *et al*., 2018).

## Study Limitations

This study had several limitations that should be considered when interpreting the findings. First, the plasma samples used in this study were not linked to detailed patient clinical information. Consequently, HBV DNA levels, identified genotypes, and detected HBsAg mutations could not be correlated with individual clinical characteristics or disease outcomes.

Second, socio-demographic information for the patients, including gender, education level, marital status, and occupation, was not available. As a result, it was not possible to assess potential associations between these factors and HBV infection patterns observed in the study population.

## Conclusion

This study revealed a significant prevalence of HBV infection among HIV-positive individuals undergoing antiretroviral therapy in Nairobi County. It highlights the ongoing public health importance of co-infection with HIV and HBV. Molecular analysis showed the presence of HBV genotypes A (sub-genotypes A1 and A4) and D (sub-genotypes D4 and D10) within the population studied. Additionally, mutations in the HBV S gene, particularly R122K and Y134F, were identified, indicating potential impacts on viral antigenicity, diagnostic detection, and immune escape. These findings emphasize the necessity for routine HBV screening among individuals living with HIV and the need for ongoing molecular surveillance of circulating HBV genotypes and mutations. Further research involving larger populations and clinical data is recommended to enhance the understanding of the epidemiological and clinical significance of HBV genetic diversity and mutation patterns in co-infected populations.

## Supporting information

Supplementary Table 1: GenBank accession numbers assigned to HBV S gene sequences generated in this study.

## Data Availability

The Hepatitis B Virus (HBV) surface (S) gene sequences generated in this study have been deposited in the National Center for Biotechnology Information (NCBI) GenBank database under accession numbers PZ062441 to PZ062474.

## Acknowledgements

The authors would like to acknowledge the staff of the KEMRI Laboratory for Molecular Biology and the University of Nairobi, Center for Biotechnology and Bioinformatics for their technical support and assistance during the laboratory work. Their contributions to sample processing, data generation, and overall research support were invaluable to the successful completion of this study.

