## Supplementary Table 1: GenBank accession numbers assigned to HBV S gene sequences generated in this study. for "Prevalence, Genotyping, and Mutational Analysis of Hepatitis B Virus in HIV-Infected Patients on Antiretroviral Therapy in Nairobi, Kenya"

### **Supplementary Materials**

Supplementary Table 1: GenBank accession numbers assigned to HBV S gene sequences generated in this study.

| **Study Sample Sequence** | **Accession Number** |
| --- | --- |
| HBV 004 | PZ062441 |
| HBV 012 | PZ062442 |
| HBV 015 | PZ062443 |
| HBV 032 | PZ062444 |
| HBV 043 | PZ062445 |
| HBV 049 | PZ062446 |
| HBV 086 | PZ062447 |
| HBV 089 | PZ062448 |
| HBV 090 | PZ062449 |
| HBV 091 | PZ062450 |
| HBV 092 | PZ062451 |
| HBV 093 | PZ062452 |
| HBV 095 | PZ062453 |
| HBV 096 | PZ062454 |
| HBV 098 | PZ062455 |
| HBV 101 | PZ062456 |
| HBV 103 | PZ062457 |
| HBV 104 | PZ062458 |
| HBV 109 | PZ062459 |
| HBV 110 | PZ062460 |
| HBV 112 | PZ062461 |
| HBV 113 | PZ062462 |
| HBV 120 | PZ062463 |
| HBV 133 | PZ062464 |
| HBV 137 | PZ062465 |
| HBV 146 | PZ062466 |
| HBV 150 | PZ062467 |
| HBV 158 | PZ062468 |
| HBV 160 | PZ062469 |
| HBV 181 | PZ062470 |
| HBV 183 | PZ062471 |
| HBV 186 | PZ062472 |
| HBV 201 | PZ062473 |
| HBV 482 | PZ062474 |
